# Association between Gestational Weight Gain on Obstetric–Perinatal Outcomes Among Women With Pre-pregnancy Overweight or Obesity in a Peruvian Public Hospital

**DOI:** 10.64898/2026.03.12.26348271

**Authors:** Francisco Cesar Hernández-Concepción, Alessandra Peña-Cano, Jorge Eduardo Dávila-Quispealaya, Katty Manrique-Franco, Wendy M. Yánac-Tellería, Marlon Yovera-Aldana

## Abstract

**Objective:** To evaluate the association between excessive gestational weight gain (GWG) and obstetric and perinatal outcomes among women with pre-pregnancy excess weight attending a public hospital in Lima, Peru.

**Methods:** We conducted a retrospective cohort study using routinely collected institutional records from Hospital María Auxiliadora. Women with singleton pregnancies and pre-pregnancy body mass index (BMI) ≥25 kg/m² who delivered between January 2024 and August 2025 were included. Excessive versus non-excessive GWG was defined according to national guidelines. The primary outcome was a composite obstetric–perinatal outcome. Crude and adjusted relative risks (RRs) were estimated using Poisson regression with robust variance. Effect modification by pre-pregnancy BMI and maternal short stature was evaluated.

**Results:** Of 6082 records, 3118 met the eligibility criteria; 31.0% had excessive GWG. In adjusted analyses, excessive GWG was associated with a small increase in the risk of the composite outcome (aRR = 1.05; 95% CI: 1.01–1.09), but not with overall obstetric outcomes (aRR = 1.04; 95% CI: 0.99–1.09) or overall perinatal outcomes (aRR = 0.99; 95% CI: 0.85–1.15). The association varied according to pre-pregnancy BMI, with higher relative risks observed among women with obesity (classes I–III).

**Conclusions:** Among women with pre-pregnancy excess weight, excessive gestational weight gain was associated with a small increase in the risk of composite obstetric–perinatal outcomes but not with obstetric or perinatal outcomes analysed separately. The magnitude of the association differed across BMI categories, with stronger associations in higher obesity classes. These findings emphasise the importance of pre-pregnancy nutritional status when interpreting the potential impact of gestational weight gain on pregnancy outcomes.

## Introduction

Maternal excess weight, a global public health challenge affecting a growing proportion of women of reproductive age, is associated with adverse maternal and child health outcomes [1]. In Peru, this trend is similarly observed, with a high prevalence of excess weight in the adult population, meaning that many women enter pregnancy with overweight or obesity[2]. An elevated pre-pregnancy Body Mass Index (BMI) has been consistently associated with increased risk of conditions such as pre-eclampsia, macrosomia and caesarean section [3].

Despite the robust evidence regarding pre-pregnancy BMI and the availability of reference guidelines for gestational weight gain, such as those issued by the Institute of Medicine (IOM), applied in Peru [4], important uncertainties remain. Although excessive gestational weight gain has been associated with an increased risk of several maternal and neonatal outcomes, including hypertensive disorders of pregnancy, gestational diabetes, caesarean delivery and neonatal risks such as macrosomia [5]. It remains unclear whether gestational weight gain confers additional risk among women who already enter pregnancy with overweight or obesity, where baseline metabolic risk is already elevated [6]. Moreover, evidence from Latin American settings is limited, particularly in Peru, where epidemiological, nutritional, and health system contexts may differ from those of populations studied in other regions [7,8].

By focusing on a local Peruvian context, this study aims to contribute evidence that may inform future prevention strategies tailored to the needs of the population. Accordingly, the objective of this study was to evaluate the association of gestational weight gain on the occurrence of obstetric and perinatal outcomes among women with pre-pregnancy excess weight receiving care at Hospital María Auxiliadora.

## METHODS

### Study design and clinical setting

A retrospective observational cohort study was conducted, based on the analysis of secondary data derived from an institutional database compiled from perinatal record booklets and obstetric clinical information systems.

The study was conducted at Hospital María Auxiliadora, a tertiary-level referral hospital within the South Lima Health Network, which predominantly serves patients affiliated with the Peruvian public health insurance system (Sistema Integral de Salud, SIS). The cohort was defined retrospectively using available records, with gestational weight gain during pregnancy considered as the exposure and obstetric or perinatal outcomes occurring at delivery or during the immediate perinatal period as the outcomes.

### Study population

The study population comprised postpartum women who delivered between January 2024 and August 2025 at Hospital María Auxiliadora, with documented records of delivery and the immediate neonatal period up to hospital discharge. Women were eligible if they had a pre-pregnancy BMI ≥25 kg/m^2^ and a singleton pregnancy. Postpartum women were excluded if there was no recorded pre-pregnancy weight, weight at the end of pregnancy, or maternal height, thereby precluding the calculation of gestational weight gain.

### Sample and sampling

The initial sample size was calculated considering the occurrence of any obstetric or perinatal outcome as the primary outcome, assuming a comparison of two proportions between women with excessive and non-excessive gestational weight gain. For this calculation, an expected outcome rate of 52% in the unexposed group was used, based on previous studies [9], and a minimum clinically relevant relative risk of 1.2 was assumed. A 95% confidence level and 80% statistical power were applied, yielding a required total sample size of 772 participants.

However, as this study was based on a secondary institutional database, no sampling procedure was undertaken. Instead, all eligible women who met the inclusion criteria and did not fulfil any exclusion criteria during the study period were included, constituting a census of all eligible women during the study period. Consequently, the final sample size was considerably larger than the minimum initially estimated. The statistical power ultimately achieved is reported in the Results section.

### Variables and operationalization

Study variables were determined by data availability and the structure of routine entries in the Perinatal Record Booklet, which influenced both variable selection and their analytical categorisation.

#### a. Dependent variables

The primary outcome was the occurrence of any obstetric or perinatal outcome [10]. Composite outcomes were used to capture the overall burden of obstetric and perinatal events and to increase statistical power given the low frequency of several individual outcomes. *Any obstetric outcome* comprised caesarean delivery, pre-eclampsia, preterm birth, postpartum haemorrhage, hysterotomy and hysterectomy. These events were identified through routine entries in the Perinatal Record Booklet and hospital discharge clinical systems at Hospital María Auxiliadora. Caesarean delivery was included in this composite outcome due to its role as an event that, although sometimes planned, it often reflects an underlying outcome or increased maternofetal risk, particularly in populations with excess weight. It represents a marker of significant maternal morbidity and increased healthcare resource utilization, consistent with the use of composite outcomes in high-impact epidemiological research[11]. We acknowledge that caesarean section does not invariably constitute an outcome per se; however, its inclusion aligns with previous studies assessing the overall burden of adverse outcomes requiring substantial obstetric intervention. *Any perinatal outcome* included macrosomia, low birth weight, intrauterine growth restriction, an Apgar score <7 at one and five minutes, admission to intermediate or intensive neonatal care, and perinatal death.

Certain outcomes were also analysed individually due to their clinical relevance, including caesarean delivery, macrosomia, and admission to the neonatal intensive care unit (NICU).

Composite outcomes were defined a priori, given the low frequency of certain individual events and the absence of a single priority outcome. Owing to the retrospective design and reliance on secondary institutional data, operational definitions were based on information recorded in the Perinatal Record Booklet and hospital discharge summaries. Consequently, standardised clinical criteria were not consistently available for all specific outcomes (e.g. postpartum haemorrhage), a limitation addressed in the relevant section.

#### b. Independent variables

Gestational weight gain was classified as excessive or non-excessive for the primary analyses according to the recommendations of the Peruvian Ministry of Health Clinical Practice Guideline [12].

For women with pre-pregnancy BMI of 25.0–29.9 kg/m^2^, weight gain was considered excessive if ≥11 kg and non-excessive if <11 kg. For those with a pre-pregnancy BMI ≥30 kg/m^2^, weight gain was defined as excessive if ≥9 kg and non-excessive if <9 kg.

For supplementary analyses, gestational weight gain was also categorised into four levels: negative gain (<0 kg); insufficient gain (<7 kg for overweight and <5 kg for obesity); adequate gain (7.0–10.99 kg for overweight and 5.0–8.99 kg for obesity); and excessive gain (as defined above).

#### c. Covariates

Covariates included maternal age (continuous and categorical; considered a potential mediator and therefore not included in the primary multivariable models), pre-pregnancy BMI (continuous and categorised as overweight, and obesity classes I, II and III), educational level (no formal/primary, secondary and higher education), marital status (with or without a partner), parity (primiparous, multiparous and grand multiparous), number of antenatal visits (<6 and ≥6), previous caesarean section (yes/no), chronic anaemia (yes/no), and gestational age at delivery (continuous and categorical). Some of these categories reflect the original structure of the clinical records rather than theoretical recoding and were retained to preserve consistency with the primary data source.

### Data collection

The database consisted of routinely collected clinical information recorded in the Perinatal Record Booklet and institutional obstetric information systems at Hospital María Auxiliadora. Following institutional approvals, the Statistics Department of Hospital María Auxiliadora provided the research team with an anonymised database compiled from these sources. The dataset was supplied in digital format and did not involve direct access to individual medical records or physical perinatal booklets. Data were evaluated and analyzed from june 1 to october 31, 2025, after approval from the hospital’s ethics committee.

The database included recorded values for pre-pregnancy weight and height, weight at the end of pregnancy, gestational age, number of antenatal visits, obstetric history, and obstetric and perinatal outcomes documented at delivery and during the immediate perinatal period.

Prior to analysis, the dataset underwent a structured data cleaning and quality control process. This included verification of plausible ranges, identification of missing and extreme values, and exclusion of records that did not meet predefined eligibility criteria (e.g. insufficient information to calculate gestational weight gain, multiple pregnancies, or pre-pregnancy body mass index < 25 kg/m²).

A complete-case approach was applied for the statistical analyses. Records with insufficient information to define the primary exposure (gestational weight gain) or the outcomes of interest were excluded. No multiple imputation procedures were performed because the proportion of missing data was low and the variables were derived from routinely recorded clinical information.

The database corresponded to routinely collected clinical information from the Perinatal Record Booklet and institutional obstetric information systems at Hospital María Auxiliadora. Following the relevant institutional approvals, the Statistics Department of Hospital María Auxiliadora provided the research team with an anonymised database compiled from the Perinatal Record Booklet and institutional obstetric clinical information systems. The dataset was supplied in digital format and did not involve direct access to individual medical records or physical perinatal booklets.

### Statistical Processing and Analysis

Statistical analyses were performed using Stata version 19.5 (StataCorp, College Station, TX, USA). A two-sided p-value < 0.05 was considered statistically significant. All confidence intervals (CIs) were calculated at the 95% level.

#### Descriptive Analysis

Baseline characteristics of the study population were described overall and stratified according to excessive gestational weight gain, defined as the primary exposure. Continuous variables were summarised as mean and standard deviation (SD) when approximately normally distributed, or as median and interquartile range (IQR; 25th–75th percentile) when the normality assumption was not met. Distributional assumptions were assessed using graphical inspection and statistical tests. Categorical variables were presented as absolute frequencies and percentages.

Between-group comparisons (excessive vs non-excessive gestational weight gain) were conducted using Student’s t-test or the Wilcoxon rank-sum test, as appropriate, for continuous variables, and the chi-squared test for categorical variables.

#### Bivariable Analysis of Outcomes

The frequency of obstetric, perinatal, and combined obstetric–perinatal outcomes was estimated according to gestational weight gain category, together with corresponding 95% CIs. Crude relative risks (RRs) were estimated using Poisson regression models with robust variance, comparing women with excessive gestational weight gain to those with non-excessive weight gain.

#### Multivariable Analysis

The primary outcome was any obstetric–perinatal outcome. Secondary outcomes included any obstetric outcome and any perinatal outcome. Individual outcomes such as caesarean section, macrosomia, and neonatal admission were analysed exploratorily. Adjusted associations were estimated using Poisson regression models with robust variance, and results were reported as adjusted relative risks (aRRs) with 95% confidence intervals (CI). This approach was used because the composite outcome was common in the study population, allowing direct estimation of relative risks and avoiding the overestimation that may occur with odds ratios from logistic regression when outcomes are common.

Covariate selection was based on clinical and epidemiological criteria and informed by a prespecified causal framework constructed using directed acyclic graphs (DAGs). Variables considered potential mediators in the causal pathway were not included in the main multivariable models to avoid overadjustment.

Based on the DAGs, maternal age, pre-pregnancy body mass index, parity, number of antenatal visits, and educational level were identified as potential confounders of the association between gestational weight gain and composite outcomes. Gestational age was considered a mediator in the causal pathway and was therefore not included as a covariate in the primary multivariable models to avoid blocking relevant causal pathways. Short maternal stature was considered a confounder only in the analysis of perinatal outcomes, given its biological plausibility in relation to fetal growth and neonatal outcomes. All analyses followed a complete-case approach, including only records with available data for all variables specified in each model.

#### Assessment of Interaction (Effect Modification)

Given that gestational weight gain recommendations are defined according to pre-pregnancy body mass index (BMI), and that short maternal stature may plausibly modify the association between weight gain and adverse outcomes, potential effect modification by both variables was formally assessed. Multiplicative interaction terms were introduced into the regression models between excessive gestational weight gain and categorised pre-pregnancy BMI, as well as between gestational weight gain and maternal height. Statistical significance of interaction terms was evaluated using Wald tests. Where evidence of interaction was observed, stratum-specific relative risks were estimated using linear combinations of model coefficients (Stata lincom command), allowing interpretation of the effect of excessive gestational weight gain within each stratum.

### Ethical Considerations

The project (PRE-15-2024-00024) was reviewed by the Institutional Review Board/Ethics Committee of Universidad Científica del Sur and approved under Certificate No. 500-CIEI-CIENTIFICA-2024. It was also reviewed and approved by the Institutional Ethics Committee of Hospital María Auxiliadora under de Unique Registration code HMA/CIEI/052/2024. Access was granted to an anonymized database that did not include names, surnames, medical record numbers, or any other personally identifiable information.

## Results

### 1. Participant Selection Flow

A total of 6082 women who delivered between January 2024 and August 2025 were initially identified. Of these, 900 were excluded due to incomplete exposure data (baseline weight, final weight, or height), leaving 5272 women with complete data to calculate gestational weight gain.

Subsequently, 2154 women were excluded for not meeting the eligibility criteria of the target population, primarily due to a pre-pregnancy BMI < 25 kg/m^2^ or multiple pregnancy. The final analytical sample comprised 3,118 postpartum women **(Figure 1).**

**Figure 1.**
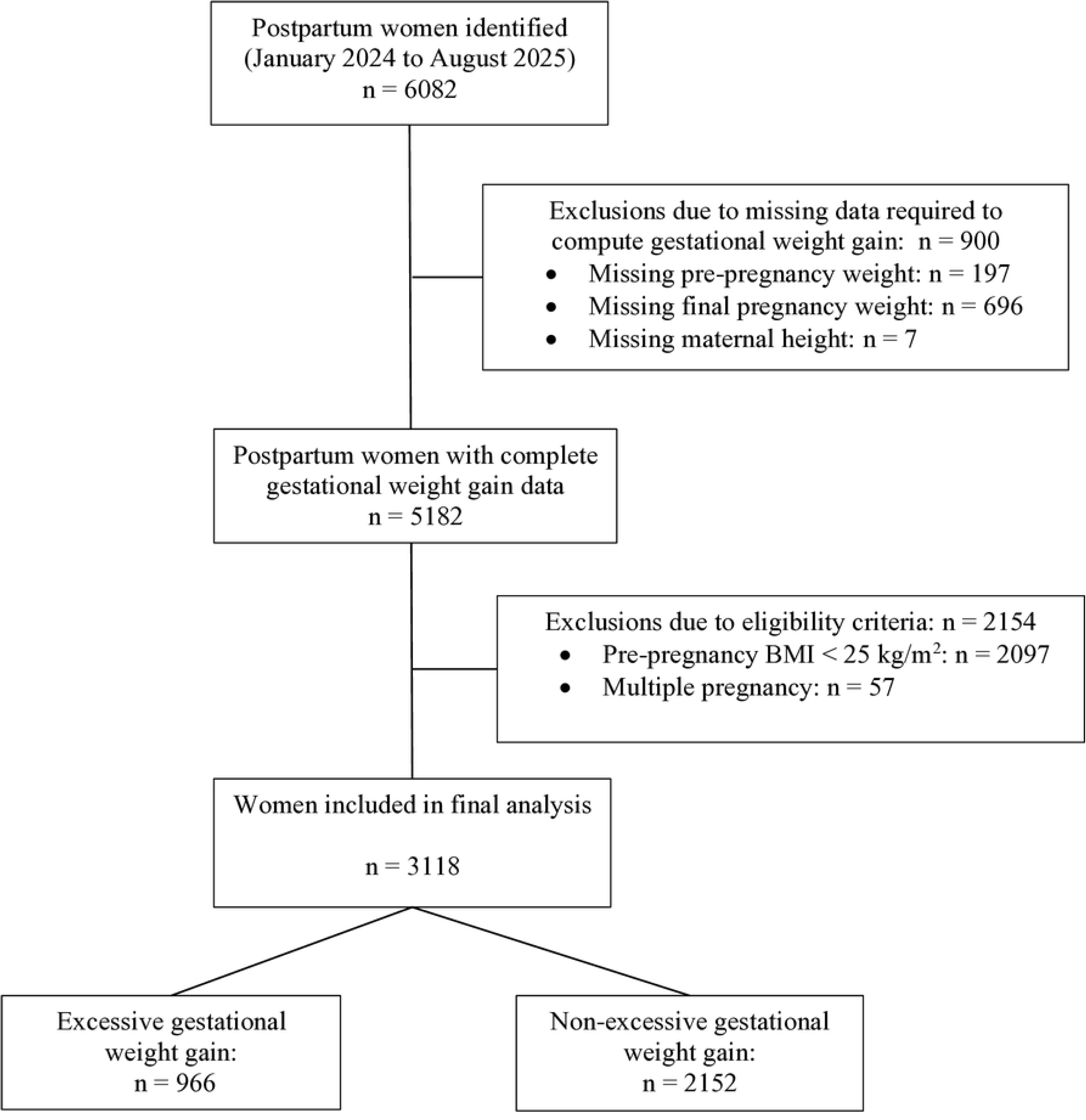
Flow diagram of participant selection. GWG was calculated as the difference between weight at the end of pregnancy and pre-pregnancy weight. Records lacking the information required to compute GWG were excluded.

Compared with those excluded, included participants tended to be older, attended a higher number of antenatal care visits, and more frequently presented with multiparity, advanced maternal age, and a history of previous caesarean section. These differences were expected with the predefined inclusion and exclusion criteria **(S1 Table)**.

Missing data were minimal for most variables, with the exception of interpregnancy interval and chronic anaemia **(S2 Table)**.

### 2. Baseline Characteristics of Study Population

The mean maternal age was 29.8 ± 6.6 years, and most women were aged 25–34 years (50.4%). Women with excessive gestational weight gain were slightly younger than those without it (28.9 vs 30.1 years; p < 0.001). According to pre-pregnancy BMI, most participants were classified as overweight (61.6%), followed by class I obesity (30.2%). No statistically significant differences were identified with respect to educational level, marital status, maternal height, interpregnancy interval, or previous caesarean section **(Table 1).**

**Table 1.** Baseline characteristics according to excessive gestational weight gain.

| Characteristic | Total (N=3,118) | Non-excessive GWG (n=2,152) | Excessive GWG (n=966) | p-value |
| --- | --- | --- | --- | --- |
| **Sociodemographic characteristics** |  |  |  |  |
| Maternal age (years), mean (SD) | 29.8 (6.6) | 30.1 (6.5) | 28.9 (6.5) | <0.001 |
| Maternal age category |  |  |  | 0.001 |
| 15.0 – 17.9 years | 69 (2.2) | 41 (1.9) | 28 (2.9) |  |
| 18.0 – 24.9 years | 674 (21.6) | 430 (20.0) | 244 (25.3) |  |
| 25.0 – 34.9 years | 1571 (50.4) | 1095 (50.9) | 476 (49.3) |  |
| 35.0 – 50.0 years | 802 (25.7) | 584 (27.1) | 218 (22.6) |  |
| Educational level |  |  |  | 0.082 |
| Illiterate / primary | 131 (4.2) | 101 (4.6) | 30 (3.1) |  |
| Secondary | 2266 (72.7) | 1546 (71.9) | 720 (74.6) |  |
| Higher education | 717 (23.0) | 502 (23.4) | 215 (22.3) |  |
| Marital status |  |  |  | 0.100 |
| With partner | 2000 (69.8) | 1403 (70.8) | 597 (67.7) |  |
| Without partner | 862 (30.1) | 578 (29.2) | 284 (32.2) |  |
| **Anthropometric characteristics** |  |  |  |  |
| Pre-pregnancy BMI ($\text{kg}/\text{m}^2$), mean (SD) | 29.6 (3.5) | 29.8 (3.6) | 28.9 (3.1) | <0.001 |
| Pre-pregnancy BMI category |  |  |  | <0.001 |
| 25.0–29.9 $\text{kg}/\text{m}^2$ | 1923 (61.6) | 1283 (59.6) | 640 (66.2) | |
| 30.0–34.9 $\text{kg}/\text{m}^2$ | 943 (30.2) | 662 (30.7) | 281 (29.0) | |
| 35.0–39.9 $\text{kg}/\text{m}^2$ | 222 (7.1) | 179 (8.3) | 43 (4.5) | |
| $\geq 40.0 \text{ kg}/\text{m}^2$ | 30 (0.9) | 28 (1.3) | 2 (0.2) | |
| Maternal stature |  |  |  | 0.279 |
| Non-short stature ($\geq 157 \text{ cm}$) | 959 (30.8) | 649 (30.1) | 310 (32.0) | |
| Short stature ($< 157 \text{ cm}$) | 2159 (69.2) | 1503 (69.8) | 656 (67.9) | |
| **Obstetric characteristics** |  |  |  |  |
| Gestational age at delivery (weeks), mean (SD) | 38.1 (2.2) | 38.0 (2.4) | 38.5 (1.7) | <0.001 |
| Gestational age |  |  |  | <0.001 |
| Preterm (<37 weeks) | 415 (13.3) | 329 (15.2) | 86 (8.8) |  |
| Term (37–41 weeks) | 2699 (86.5) | 1821 (84.34) | 878 (90.1) |  |
| Post-term (≥42 weeks) | 4 (0.1) | 2 (0.1) | 2 (0.2) |  |
| Antenatal visits, mean (SD) | 7.1 (2.0) | 7.0 (2.1) | 7.4 (1.9) | <0.001 |
| N° of antenatal visits |  |  |  | <0.001 |
| <6 | 667 (21.3) | 499 (23.2) | 168 (17.3) |  |
| ≥6 | 2,451 (78.6) | 1653 (76.8) | 798 (82.6) |  |
| Interpregnancy interval, (months) median [IQR] | 57.5 [29.4–95.5] | 57.3 [28.1–96.3] | 58.6 [31.2–91.9] | 0.851 |
| Gravidity |  |  |  | <0.001 |
| Primigravida | 685 (21.9) | 439 (20.3) | 246 (25.5) |  |
| Multigravida (2–3) | 1604 (51.4) | 1102 (51.2) | 502 (51.9) |  |
| Grand multigravida (≥4) | 829 (26.6) | 611 (28.4) | 218 (22.6) |  |
| Parity |  |  |  | <0.001 |
| Nulliparous | 920 (29.5) | 591 (27.4) | 329 (34.1) |  |
| Multiparous (1–2) | 1756 (56.3) | 1233 (57.3) | 523 (54.1) |  |
| Grand multiparous (≥3) | 442 (14.1) | 328 (15.2) | 114 (11.8) |  |
| Previous caesarean section |  |  |  | 0.456 |
| No | 2435 (93.1) | 1672 (92.9) | 763 (93.7) |  |
| Yes | 178 (6.8) | 127 (7.1) | 51 (6.3) |  |
| Chronic anaemia, n=1863 |  |  |  | 0.014 |
| No | 911 (72.5) | 604 (70.5) | 307 (77.1) |  |
| Yes | 344 (27.4) | 253 (29.5) | 91 (22.9) |  |

GWG, gestational weight gain; SD, standard deviation; IQR, interquartile range.
Continuous variables are presented as mean (SD) or median [IQR], according to
distribution. Categorical variables are presented as n (%). Group comparisons were
performed using Student's t test, Wilcoxon rank-sum test, or chi-square test, as
appropriate. P-values <0.001 are reported as such.

Regarding obstetric characteristics, women with excessive gestational weight gain had a higher gestational age at delivery (38.5 vs 38.0 weeks; p < 0.001), attended a greater number of antenatal care visits on average (7.4 vs 7.0; p < 0.001), and more frequently attended ≥6 antenatal visits (32.6% vs 25.2%). Chronic anaemia was also more prevalent in this group (33.7% vs 25.6%; p = 0.014) **(Table 1).**

Overall, 31.0% of women experienced excessive gestational weight gain (95% CI: 29.3–32.6). It was highest among women with overweight (33.3%) and decreased across obesity classes I, II, and III. This pattern was consistent among women with short stature and those without short stature **(Table 2).**

**Table 2.**
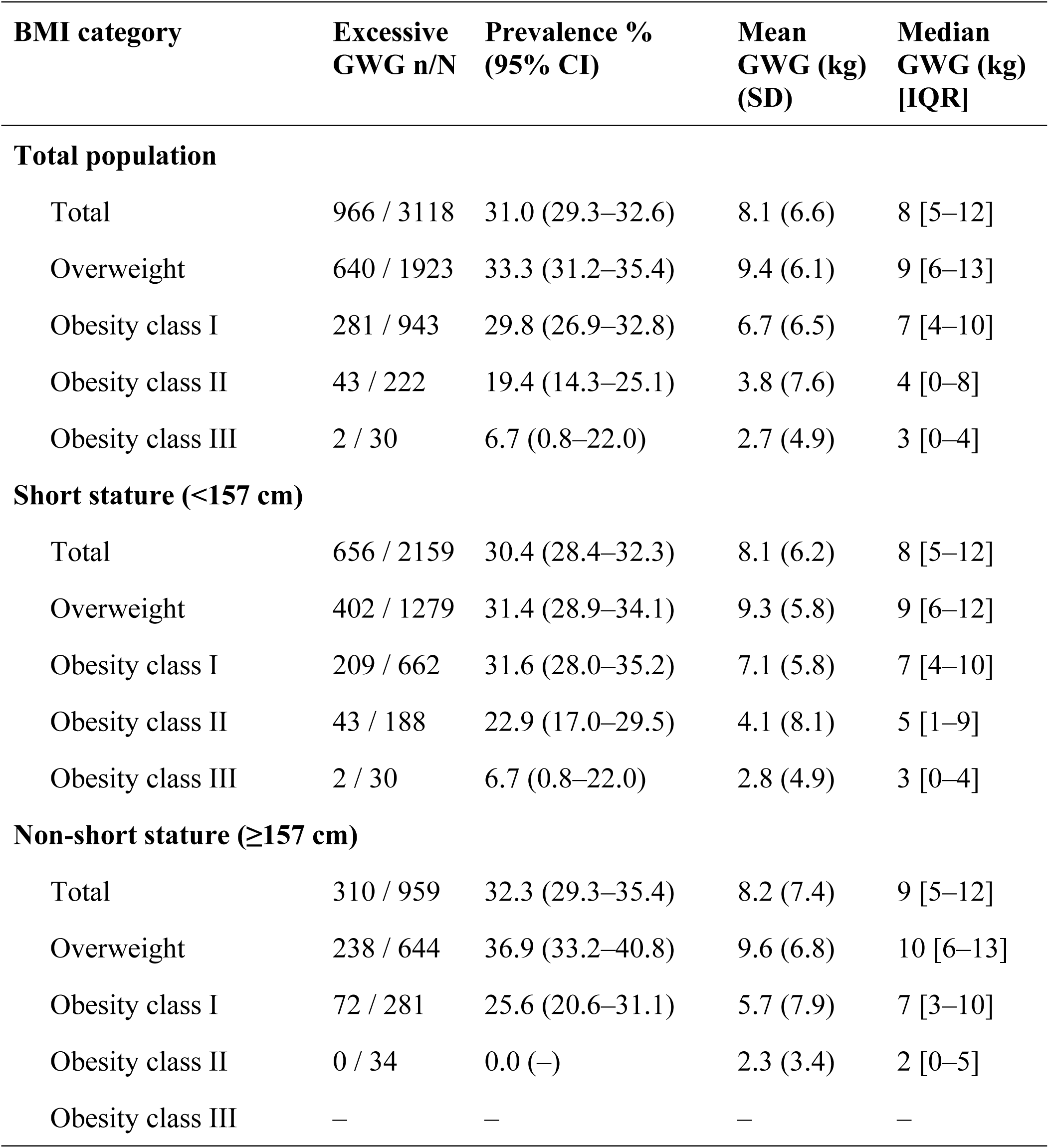
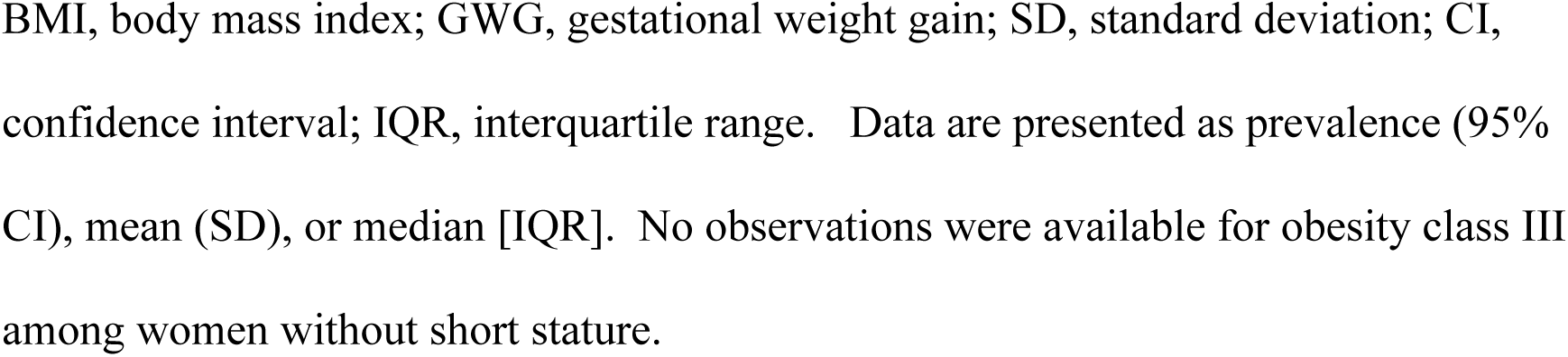
Prevalence of excessive gestational weight gain by pre-pregnancy BMI and maternal height.

| BMI category | Excessive GWG n/N | Prevalence % (95% CI) | Mean GWG (kg) (SD) | Median GWG (kg) [IQR] |
| --- | --- | --- | --- | --- |
| **Total population** |  |  |  |  |
| Total | 966 / 3118 | 31.0 (29.3–32.6) | 8.1 (6.6) | 8 [5–12] |
| Overweight | 640 / 1923 | 33.3 (31.2–35.4) | 9.4 (6.1) | 9 [6–13] |
| Obesity class I | 281 / 943 | 29.8 (26.9–32.8) | 6.7 (6.5) | 7 [4–10] |
| Obesity class II | 43 / 222 | 19.4 (14.3–25.1) | 3.8 (7.6) | 4 [0–8] |
| Obesity class III | 2 / 30 | 6.7 (0.8–22.0) | 2.7 (4.9) | 3 [0–4] |
| **Short stature (<157 cm)** |  |  |  |  |
| Total | 656 / 2159 | 30.4 (28.4–32.3) | 8.1 (6.2) | 8 [5–12] |
| Overweight | 402 / 1279 | 31.4 (28.9–34.1) | 9.3 (5.8) | 9 [6–12] |
| Obesity class I | 209 / 662 | 31.6 (28.0–35.2) | 7.1 (5.8) | 7 [4–10] |
| Obesity class II | 43 / 188 | 22.9 (17.0–29.5) | 4.1 (8.1) | 5 [1–9] |
| Obesity class III | 2 / 30 | 6.7 (0.8–22.0) | 2.8 (4.9) | 3 [0–4] |
| **Non-short stature (≥157 cm)** |  |  |  |  |
| Total | 310 / 959 | 32.3 (29.3–35.4) | 8.2 (7.4) | 9 [5–12] |
| Overweight | 238 / 644 | 36.9 (33.2–40.8) | 9.6 (6.8) | 10 [6–13] |
| Obesity class I | 72 / 281 | 25.6 (20.6–31.1) | 5.7 (7.9) | 7 [3–10] |
| Obesity class II | 0 / 34 | 0.0 (–) | 2.3 (3.4) | 2 [0–5] |
| Obesity class III | – | – | – | – |

BMI, body mass index; GWG, gestational weight gain; SD, standard deviation; CI, confidence interval; IQR, interquartile range. Data are presented as prevalence (95% CI), mean (SD), or median [IQR]. No observations were available for obesity class III among women without short stature.

A detailed distribution of non-excessive gestational weight gain (negative, insufficient, and adequate) according to National Institute of Health criteria is presented in **S3 Table.**.

### 3. Frecuency of Maternal and Perinatal Outcomes

In the crude analysis, excessive gestational weight gain was associated with a higher frequency of any obstetric–perinatal outcome compared with non-excessive weight gain (78.9% vs 75.9%), corresponding to a smalll increase in risk (RR = 1.04; 95% CI: 1.00–1.08; p = 0.060). No statistically significant associations were observed for overall obstetric outcomes (RR = 1.03; 95% CI: 0.99–1.08; p = 0.139) or overall perinatal outcomes (RR = 0.93; 95% CI: 0.81–1.08; p = 0.369).

Among specific outcomes, excessive gestational weight gain was associated with an increased risk of caesarean section (RR = 1.13; 95% CI: 1.06–1.20) and macrosomia (RR = 1.76; 95% CI: 1.40–2.22).

Conversely, it was associated with lower risks of preterm birth (RR = 0.58; 95% CI: 0.47–0.73), low birth weight (RR = 0.43; 95% CI: 0.30–0.61), and intrauterine growth restriction (RR = 0.58; 95% CI: 0.40–0.85). Lower risks were also observed for Apgar score <7 at 1 minute (RR = 0.66; 95% CI: 0.44–0.99) and neonatal admission (RR = 0.57; 95% CI: 0.41–0.79).

No significant associations were identified with pre-eclampsia, hysterectomy, Apgar score <7 at 5 minutes, or perinatal mortality (**Table 3** and **S1 Figure**).

**Table 3.** Frequency and crude relative risks of obstetric and perinatal outcomes according to gestational weight gain among women with pre-pregnancy excess weight.

| Outcomes | Excessive GWG (N = 966) | Non-excessive GWG (N = 2,152) | Crude RR (95% CI) | p-value |
| --- | --- | --- | --- | --- |
| **Any obstetric–perinatal outcome** | 762 (78.9) | 1633 (75.9) | 1.04 (1.00–1.08) | 0.060 |
| **Obstetric outcomes** | 730 (75.6) | 1573 (73.1) | 1.03 (0.99–1.08) | 0.139 |
| Preeclampsia | 42 (4.4) | 75 (3.5) | 1.25 (0.86–1.81) | 0.242 |
| Preterm birth | 86 (8.9) | 329 (15.3) | 0.58 (0.47–0.73) | <0.001 |
| Caesarean section | 590 (61.1) | 1168 (54.3) | 1.13 (1.06–1.20) | <0.001 |
| Postpartum haemorrhage | 707 (73.2) | 1501 (69.8) | 1.05 (1.00–1.10) | 0.046 |
| Hysterectomy | 1 (0.1) | 2 (0.1) | 1.11 (0.10–12.27) | 0.930 |
| Hysterotomy (<30 weeks) | 0 (0.0) | 20 (0.9) | NC |  |
| **Perinatal outcomes** | 198 (20.5) | 472 (21.9) | 0.93 (0.81–1.08) | 0.369 |
| Macrosomia | 116 (12.0) | 147 (6.8) | 1.76 (1.40–2.22) | <0.001 |
| Low birth weight | 34 (3.5) | 178 (8.3) | 0.43 (0.30–0.61) | <0.001 |
| Intrauterine growth restriction | 32 (3.3) | 123 (5.7) | 0.58 (0.40–0.85) | 0.005 |
| Apgar score <7 at 1 minute | 29 (3.0) | 98 (4.6) | 0.66 (0.44–0.99) | 0.043 |
| Apgar score <7 at 5 minutes | 2 (0.2) | 17 (0.8) | 0.26 (0.06–1.12) | 0.072 |
| Admission to intermediate or intensive care | 42 (4.4) | 164 (7.6) | 0.57 (0.41–0.79) | 0.001 |
| Perinatal death | 7 (0.7) | 24 (1.1) | 0.65 (0.28–1.50) | 0.314 |

GWG, gestational weight gain; RR, relative risk; CI, confidence interval. Data are presented as n (%) or crude relative risks (RR) with 95% confidence intervals (CI). Crude RRs were estimated using Poisson regression with robust variance. NC indicates that the RR could not be estimated due to absence of events in one group.

### 4. Adjusted Analysis of the Effect of Gestational Weight Gain

In the multivariable analysis, excessive gestational weight gain was independently associated with a 5% higher risk of any obstetric–perinatal outcome (aRR= 1.05; 95% CI: 1.01–1.09), after adjustment for maternal age, pre-pregnancy BMI, parity, antenatal care visits, and educational level.

For overall obstetric outcomes, excessive weight gain showed a non-significant trend towards increased risk (aRR = 1.04; 95% CI: 0.99–1.09), adjusting for the same covariates. In contrast, no association was observed between excessive gestational weight gain and overall perinatal outcomes (aRR = 0.99; 95% CI: 0.85–1.15), adjusted for the same covariates (**Table 4** y **S2 Figure**).

**Table 4.**
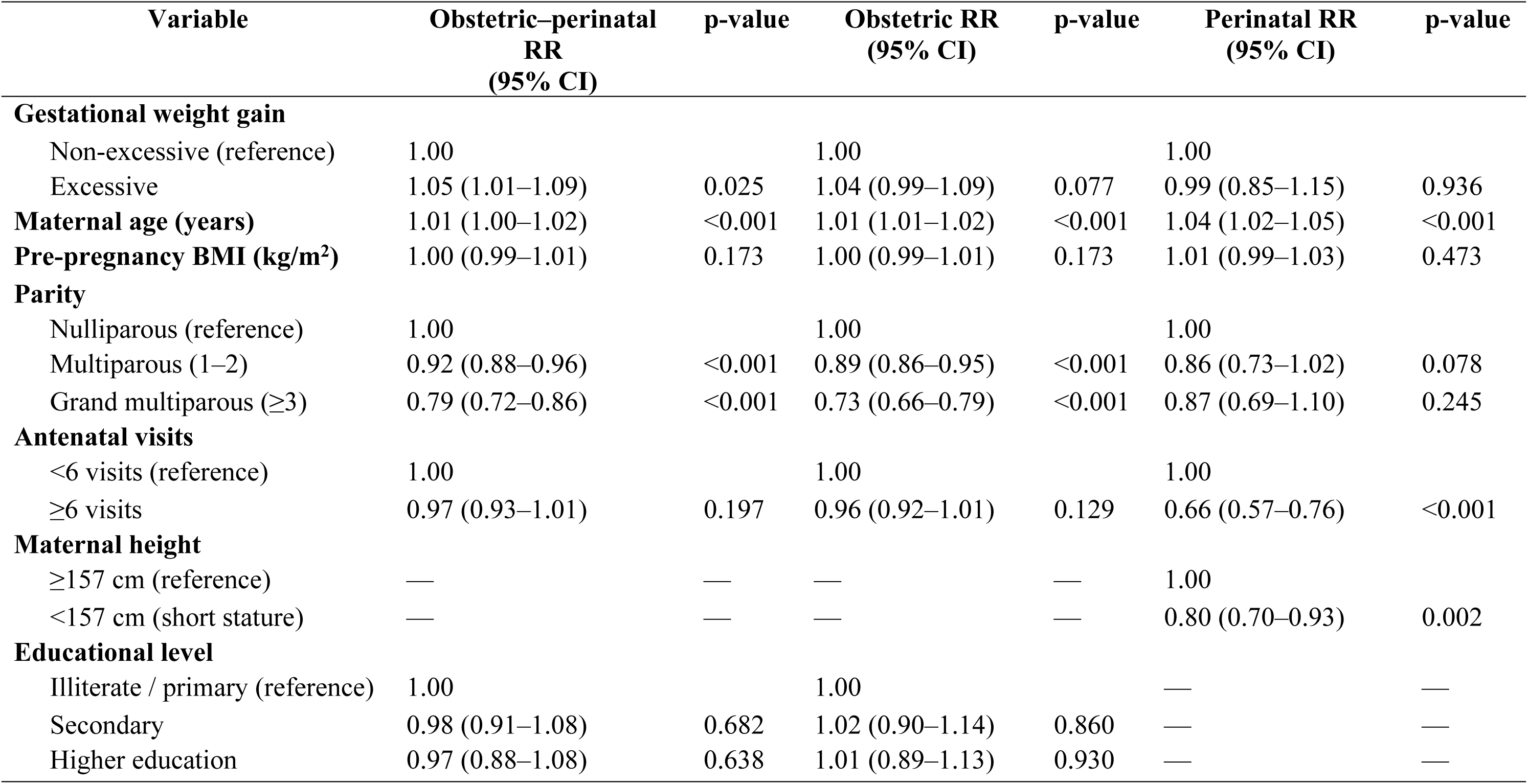
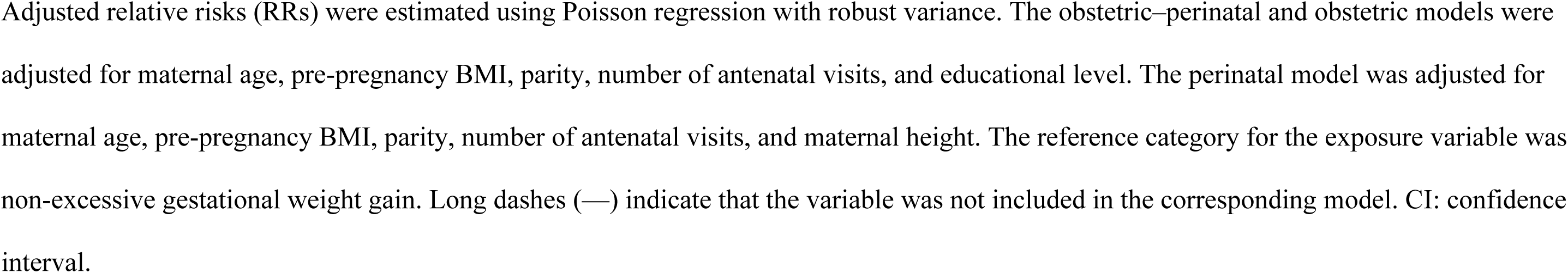
Adjusted relative risks for obstetric–perinatal, obstetric, and perinatal outcomes associated with excessive gestational weight gain among women with pre-pregnancy excess weight (N=3112, complete cases)

| Variable | Obstetric–perinatal<br>RR<br>(95% CI) | p-value | Obstetric RR<br>(95% CI) | p-value | Perinatal RR<br>(95% CI) | p-value |
| --- | --- | --- | --- | --- | --- | --- |
| **Gestational weight gain** |  |  |  |  |  |  |
| Non-excessive (reference) | 1.00 |  | 1.00 |  | 1.00 |  |
| Excessive | 1.05 (1.01–1.09) | 0.025 | 1.04 (0.99–1.09) | 0.077 | 0.99 (0.85–1.15) | 0.936 |
| **Maternal age (years)** | 1.01 (1.00–1.02) | <0.001 | 1.01 (1.01–1.02) | <0.001 | 1.04 (1.02–1.05) | <0.001 |
| **Pre-pregnancy BMI (kg/m<sup>2</sup>)** | 1.00 (0.99–1.01) | 0.173 | 1.00 (0.99–1.01) | 0.173 | 1.01 (0.99–1.03) | 0.473 |
| **Parity** |  |  |  |  |  |  |
| Nulliparous (reference) | 1.00 |  | 1.00 |  | 1.00 |  |
| Multiparous (1–2) | 0.92 (0.88–0.96) | <0.001 | 0.89 (0.86–0.95) | <0.001 | 0.86 (0.73–1.02) | 0.078 |
| Grand multiparous (≥3) | 0.79 (0.72–0.86) | <0.001 | 0.73 (0.66–0.79) | <0.001 | 0.87 (0.69–1.10) | 0.245 |
| **Antenatal visits** |  |  |  |  |  |  |
| <6 visits (reference) | 1.00 |  | 1.00 |  | 1.00 |  |
| ≥6 visits | 0.97 (0.93–1.01) | 0.197 | 0.96 (0.92–1.01) | 0.129 | 0.66 (0.57–0.76) | <0.001 |
| **Maternal height** |  |  |  |  |  |  |
| ≥157 cm (reference) | — | — | — | — | 1.00 |  |
| <157 cm (short stature) | — | — | — | — | 0.80 (0.70–0.93) | 0.002 |
| **Educational level** |  |  |  |  |  |  |
| Illiterate / primary (reference) | 1.00 |  | 1.00 |  | — | — |
| Secondary | 0.98 (0.91–1.08) | 0.682 | 1.02 (0.90–1.14) | 0.860 | — | — |
| Higher education | 0.97 (0.88–1.08) | 0.638 | 1.01 (0.89–1.13) | 0.930 | — | — |

Adjusted relative risks (RRs) were estimated using Poisson regression with robust variance. The obstetric–perinatal and obstetric models were adjusted for maternal age, pre-pregnancy BMI, parity, number of antenatal visits, and educational level. The perinatal model was adjusted for maternal age, pre-pregnancy BMI, parity, number of antenatal visits, and maternal height. The reference category for the exposure variable was non-excessive gestational weight gain. Long dashes (—) indicate that the variable was not included in the corresponding model. CI: confidence interval.

In exploratory analyses of individual outcomes, excessive gestational weight gain remained consistently associated with an increased risk of caesarean section (aRR = 1.16; 95% CI: 1.08–1.24) and macrosomia (aRR = 1.69; 95% CI: 1.35–2.13), after adjustment for maternal, obstetric, and antenatal care variables. No association was observed between excessive weight gain and neonatal admission to intermediate or intensive care (aRR = 0.97; 95% CI: 0.70–1.36) **(S4 Table)**.

### 5. Effect Modification by Pre-pregnancy BMI and Short Maternal Stature

Exploratory analyses assessed potential effect modification by pre-pregnancy BMI and short maternal stature, given that gestational weight gain recommendations are defined according to BMI and that maternal height may influence the clinical interpretation of weight gain.

The association between excessive gestational weight gain and any obstetric–perinatal outcome varied across categories of pre-pregnancy BMI, with higher relative risk estimates observed in higher BMI strata (p for interaction <0.05). No evidence of effect modification by maternal height was observed **(S5 Table)**.

## Discussion

### Main Findings

In this retrospective study of 3118 women with pre-pregnancy overweight or obesity, 31.0% experienced excessive gestational weight gain. After multivariable adjustment, excessive weight gain was associated with a 5% increased risk of any obstetric–perinatal outcome. Although statistically significant, the magnitude of this association was modest, suggesting a limited independent contribution of excessive gestational weight gain to overall obstetric–perinatal risk. No associations were observed with obstetric or perinatal outcomes considered separately. In exploratory analyses of individual outcomes, excessive gestational weight gain was associated with a 16% higher risk of caesarean section and a 69% higher risk of macrosomia.

Importantly, the association between excessive gestational weight gain and any obstetric–perinatal outcome became progressively stronger across increasing categories of pre-pregnancy BMI. This finding suggests that the clinical impact of the observed increase in risk may be greater among women with more severe pre-pregnancy obesity, in whom excessive weight gain may compound an already elevated baseline risk.

### Comparison with Previous Studies

The use of composite outcomes has been widely adopted in the literature to capture the overall burden of obstetric and neonatal events associated with maternal overweight and obesity. In this context, Soto-Sánchez *et al.*[9] reported a 51.5% prevalence of outcomes among women with obesity using a composite obstetric–perinatal outcome that integrated multiple events. However, their cross-sectional design limited the ability to assess temporal relationships or causal inference. Similarly, the meta-analysis by Goldstein *et al.*[11], which predominantly included populations from North America, Europe, and Asia, demonstrated that excessive gestational weight gain was associated with modest but consistent increases in the risk of composite maternal and neonatal outcomes, particularly caesarean section and macrosomia, with odds ratios ranging from 1.3 to 1.95. Evidence from low- and middle-income countries (LMICs) has shown comparable patterns. For instance, the meta-analysis by Darling *et al.* [13] found that excessive gestational weight gain was associated with an increased risk of large-for-gestational-age infants, macrosomia, and caesarean delivery. In addition, Perumal *et al.*[14] reported associations between suboptimal gestational weight gain and adverse neonatal outcomes in LMICs. Overall, these international findings are consistent with the present study, which observed a modest increase in the risk of composite obstetric–perinatal outcomes associated with excessive gestational weight gain.

When individual outcomes were examined, the association between excessive gestational weight gain and increased risks of caesarean section and macrosomia was consistent. Recent evidence from high-income settings supports these findings. For example, Benhalima *et al.* in Belgium [15], reported that excessive weight gain among women with obesity was associated with a significantly higher incidence of emergency caesarean section and macrosomia. Similarly, in low- and middle-income settings, a study conducted in Thailand [16] found that women with overweight or obesity who experienced excessive gestational weight gain had higher odds of caesarean delivery (adjusted OR 1.65) and macrosomia (adjusted OR 5.84). In China, a study among women with gestational diabetes [17] also demonstrated that excessive weight gain in the overweight/obese group increased the risk of macrosomia and caesarean delivery. These findings are consistent with those of the present study, in which excessive gestational weight gain was associated with a 16% higher risk of caesarean section and a 69% higher risk of macrosomia.

In the Latin American context, the available evidence also supports these associations, although with some particular nuances. The Birth in Brazil study[18] reported that gestational weight gain exceeding twice the recommended amount was associated with an increased risk of caesarean section among women with normal BMI, overweight, and obesity; however, the magnitude of the effect diminished and was no longer statistically significant in more severe obesity categories. In Peru, previous studies have examined gestational weight gain and related outcomes. Apaza Valencia *et al.*[19] described a progressive increase in macrosomia with greater gestational weight gain among women in Arequipa, while Magallanes-Corimanya *et al.* [20] reported a high frequency of excessive gestational weight gain among Peruvian women with overweight and obesity, associated with higher neonatal birth weight. Although heterogeneous in methodology, these national studies reinforce the importance of monitoring gestational weight gain in the region.

Importantly, the interaction with pre-pregnancy BMI must be considered. Findings from studies such as that by Lubrano *et al.* [21], conducted in populations with extreme pre-pregnancy obesity, suggest that excessive gestational weight gain may not be independently associated with major outcomes after multivariable adjustment, with pre-pregnancy BMI emerging as a major determinant. This highlights the complexity of the relationship and aligns with the interaction observed in the present study, in which the association between excessive gestational weight gain and any obstetric–perinatal outcome became more pronounced across increasing BMI categories.

Taken together, these findings reinforce the importance of considering pre-pregnancy BMI when interpreting the effects of gestational weight gain.

### Biological Plausibility and Underlying Mechanisms

From a pathophysiological perspective, pre-pregnancy obesity is a major determinant of obstetric and perinatal risk [3,22–26]. It establishes a maternal milieu characterised by insulin resistance, endothelial dysfunction, hormonal alterations, and chronic low-grade inflammation [27–29]. These intrinsic mechanisms increase the likelihood of obstetric outcomes independently of gestational weight gain [27]. In addition, maternal adipose tissue acts as an endocrine organ that releases adipokines such as adiponectin; reduced adiponectin levels in women with obesity have been associated with placental dysfunction and altered nutrient transport to the fetus, potentially influencing fetal growth and neonatal outcomes [28].

Within this framework, excessive gestational weight gain appears to act primarily as a modulating or exacerbating factor, amplifying pre-existing risks—particularly in women with more severe pre-pregnancy obesity [29,30]. Our findings, showing higher relative risks of composite obstetric–perinatal outcomes across increasing BMI categories, are consistent with this interaction between baseline maternal metabolic status and gestational exposures.

Excessive gestational weight gain has been consistently associated with higher risks of caesarean section, macrosomia, and large-for-gestational-age infants, often demonstrating a dose–response pattern [11,35]. Conversely, it has not associated with a reduction in preterm birth, low birth weight, or intrauterine growth restriction. Lower frequency of these outcomes has been described, likely mediated by longer gestational duration and increased fetal growth, effectively shifting risk towards the upper end of the birth weight spectrum [11,36,37]

Accordingly, risk estimates below unity for outcomes, such as preterm birth and low birth weight, should not be interpreted as evidence of a protective effect of excessive weight gain. Rather, they reflect a redistribution of perinatal risk. The meta-analysis by Goldstein et al. [11] clearly illustrates this phenomenon, reporting lower risks of small-for-gestational-age infants and preterm birth consistent increases in macrosomia and large-for-gestational-age infants.

The reported frequency of postpartum haemorrhage, including in our study, may be influenced by heterogeneity in clinical definitions and recording practices [11]. Broader definitions based on estimated blood loss or therapeutic interventions tend to increase the reported frequency of this outcome and may partly explain weak or inconsistent associations after multivariable adjustment. The high frequency of caesarean section observed in our cohort (>54%) is consistent with global evidence demonstrating a stable, moderate association between excessive gestational weight gain and caesarean delivery [11,18,38]. The inclusion of caesarean section within the composite outcome of “any obstetric outcome” is justified because, although not invariably an outcome per se, it constitutes a major obstetric intervention [39] and a indicator of maternal morbidity [40,41].

Nevertheless, the elevated caesarean rate cannot be attributed solely to pre-pregnancy BMI or gestational weight gain. Global caesarean rates are rising beyond medically justified levels [42], a trend also observed in public hospitals in low- and middle-income countries[43]. Institutional policies, clinical practice patterns, surgical thresholds, and socio-cultural or economic factorswhich may shape delivery mode independently of maternal weight. [44,45]

Finally, short maternal stature has been more consistently associated with outcomes related to fetal growth restriction, such as higher frequencies of small-for-gestational-age infants [46]. However, evidence suggests a limited independent contribution to caesarean risk after adjustment for obstetric factors [47]. In this context, the stronger association observed between excessive gestational weight gain and composite outcomes across increasing BMI categories reinforces the interaction between baseline maternal characteristics and gestational weight dynamics.

### Public Health Implications for Hospital María Auxiliadora

The findings of this study may have implications for public health strategies and clinical practice at Hospital María Auxiliadora. Excessive gestational weight gain was associated with a 5% increase in the risk of the composite obstetric–perinatal outcome. Although this effect size is modest at the individual level, its potential public health relevance becomes more evident in populations where overweight and obesity are highly prevalent among women of reproductive age.

Hospital María Auxiliadora operates in an epidemiological context characterised by a high burden of excess weight among women of reproductive age. National data indicate that overweight and obesity affect a substantial proportion of the adult female population in Peru [48], underscoring the importance of strengthening preventive strategies targeting maternal nutrition before and during pregnancy.

Our findings suggest that addressing pre-pregnancy nutritional status may be an important component of strategies aimed at reducing obstetric–perinatal outcomes [49]. In practice, these strategies could include strengthening routine screening and referral pathways for women with overweight or obesity during antenatal care [50,51]; incorporating simple visual alerts in perinatal records or electronic medical systems to facilitate monitoring of gestational weight gain [52]; promoting peer-support or group-based educational activities that encourage healthy behaviors during pregnancy [53,54]; developing accessible educational materials on recommended gestational weight gain, balanced nutrition, and physical activity [55]; and reinforcing consistent institutional messaging through brief staff training and awareness initiatives [56]. Collectively, these measures may contribute to improved monitoring of maternal nutritional status and support healthier gestational weight trajectories. Moreover, they hold potential for cost-effectiveness by reducing the downstream burden of managing complex obstetric and neonatal outcomes associated with excess maternal weight [57].

### Research recommendations

Future studies should further explore non-excessive gestational weight gain as a heterogeneous exposure. It is important to disaggregate negative, insufficient, and adequate weight gain categories, as presented in the supplementary table of this study. Independent analyses of these subcategories are needed, since insufficient weight gain has been associated with increased risks of low birth weight, small-for-gestational-age infants, and preterm birth [14]. Grouping these categories together may obscure specific associations and differential effects on obstetric and perinatal outcomes.

Prospective studies with detailed longitudinal measurements are also needed. This could include trimester-specific assessment of gestational weight gain, as the timing of weight gain during pregnancy may exert distinct effects on maternal and fetal outcomes [58]. In addition, evaluation of maternal body composition, including fat mass and fat-free mass, should be prioritized. Total body weight does not capture the relative contribution of these components, and excess adiposity is more closely linked to cardiometabolic risk and adverse neonatal outcomes [59]. Finally, research should extend to the long-term assessment of maternal and offspring outcomes. Maternal obesity and gestational weight gain have enduring implications for both mother and child, including increased risks of obesity and chronic diseases during childhood and adulthood [60].

### Limitations and Strengths

The main limitations of this study include its retrospective design and reliance on secondary clinical records. This methodological approach may introduce measurement error and variability in data quality, as information is collected for clinical rather than research purposes [61]. Medical record completeness and accuracy can vary considerably, and data entry errors or lack of standardization in documentation are common challenges in studies based on routine clinical record [62]. Operational definitions of obstetric and perinatal outcomes were based on information recorded in perinatal records and hospital discharge summaries. Consequently, standardized clinical criteria were not always available for all specific outcomes, such as precise quantification of blood loss in postpartum hemorrhage. Variability in definitions and recording practices may therefore have influenced reported frequencies and the magnitude of associations and should be considered when interpreting the findings. In addition, incomplete or inconsistent recording of some anthropometric variables, including pre-pregnancy weight and maternal height in mall proportion of records, may have affected the completeness of the dataset. Furthermore, restricting the analysis to women with pre-pregnancy overweight or obesity limits the generalizability of the findings to women with normal body mass index.

Despite these limitations, the study has several notable strengths. The large sample size of 3118 postpartum women enhances the precision of risk estimates and allows detection of associations even for less frequent outcomes [62]. The use of multivariable models with robust estimators strengthens control for potential confounding and robustness the validity of parameter estimates [63]. Furthermore, assessment of effect modification by pre-pregnancy BMI and the availability of detailed exploratory analyses provide greater methodological depth and clinical relevance, improving understanding of the interaction between gestational weight gain and baseline nutritional status.

## Conclusiones

Among women with pre-pregnancy excess weight receiving care at Hospital María Auxiliadora, excessive gestational weight gain was associated with an overall increase in the risk of composite obstetric–perinatal outcomes, although the magnitude of this association was modest when considered in isolation. No significant associations were observed when obstetric and perinatal outcomes were analysed separately as global categories. Importantly, the effect varied according to the degree of pre-pregnancy obesity. A progressively higher risk was observed among women with moderate and severe obesity, suggesting that excessive weight gain may act as a modulating factor that amplifies an already elevated baseline risk. These findings reinforce the role of pre-pregnancy nutritional status as a key determinant of obstetric and perinatal risk and highlight the importance of strategies aimed at improving maternal nutritional status before conception and during pregnancy.

## Supplementary material

S1 Fig. Crude relative risk of maternal and perinatal outcomes associated with excessive gestational weight gain

S2 Fig. Directed acyclic graphs (DAGs) representing the hypothesised causal structure between gestational weight gain and study outcomes

S1 Table. Comparison of baseline characteristics between women included and not included in the analysis.

S2 Table. Missing data across study variables (N=3,118)

S3 Table. Distribution of gestational weight gain according to pre-pregnancy BMI

S4 Table. Adjusted relative risks of obstetric and perinatal outcomes associated with excessive gestational weight gain among women with pre-pregnancy overweight or obesity.

S5 Table. Effect modification of excessive gestational weight gain according to pre-pregnancy BMI and maternal height on obstetric and perinatal outcomes

S1 File. Analysis dataset.

## Data Availability

All relevant data are within the manuscript and its Supporting Information files

## Notes

### Competing Interest Statement

The authors have declared no competing interest.

